# Patterns of Therapeutic Adherence to Antihypertensive Treatment in a Public Mexican Hospital: A Cross-Sectional Descriptive Study

**DOI:** 10.1101/2025.07.17.25331620

**Authors:** M.G. Mares Pérez, Y. De la Cruz Ramírez, David A. Franco-Torres, Agustín Avalos, Mauricio Sánchez-Barajas

**Affiliations:** School of Medicine, Universidad La Salle Bajío, Léon, Guanajuato, Mexico; General and Family Medicine Hospital No. 21 of the Mexican Social Security Institute (IMSS), Léon, Guanajuato, Mexico

**Keywords:** Hypertension, therapeutic adherence, antihypertensive treatment, medication compliance, public health, Mexico

## Abstract

Hypertension is a major cardiovascular risk factor with high prevalence in Mexico. Therapeutic adherence is crucial for blood pressure control, yet non-adherence remains a significant challenge. This cross-sectional study evaluated antihypertensive therapeutic adherence patterns among 129 adults at General Hospital of Zone and Family Medicine Clinic No. 21 (IMSS) in Léon, Mexico. Using structured interviews and a validated instrument, we assessed sociodemographic factors, clinical parameters, and adherence levels categorized as Total, Partial, or Nonadherence. Results revealed suboptimal adherence in 62% of participants, with significant associations between adherence levels and age cohorts (p*<*0.05). These findings highlight the need for targeted interventions to improve adherence in hypertensive populations.

## 1 Introduction

Cardiovascular diseases (CVDs) constitute the preeminent cause of mortality across the Americas, concurrently representing a significant source of disability, premature death, and substantial economic burden for prevention and control programs [14]. These conditions exhibit universal prevalence, indiscriminately affecting diverse demographic strata irrespective of socioeconomic or cultural status, thereby presenting a formidable challenge to public health infrastructure within the region [1].

Hypertension (HTN), defined as a persistent elevation of systolic blood pressure (SBP) *≥*140 mm Hg and/or diastolic blood pressure (DBP) *≥*90 mm Hg in adults *≥*18 years (per the Seventh Report of the Joint National Committee on Prevention, Detection, Evaluation, and Treatment of High Blood Pressure), is a chronic pathophysiological state [1, 2]. Optimal SBP and DBP levels are critical for the efficient function of vital organs—including the heart, brain, and kidneys—and for the maintenance of overall health [2]. Globally, HTN is the predominant contributor to CVD-related mortality, implicated in 45% of cardiac deaths and 51% of stroke fatalities [4].

HTN is frequently asymptomatic; when manifestations occur (e.g., cephalalgia, dyspnea, vertigo, epistaxis, or visual disturbances), they are non-specific. Severe hypertension may induce fatigue, nausea, confusion, or tremors. Definitive diagnosis necessitates clinical blood pressure measurement—a rapid, non-invasive procedure— underscoring the imperative for professional medical assessment to evaluate associated risks [2, 4].

Therapeutic non-adherence in chronic disease management, particularly concerning hypertensive disorders, has been identified as a primary factor impeding disease control and exacerbating long-term complications [8]. While precise adherence rates remain elusive, epidemiological studies indicate that approximately 50% of hypertensive patients demonstrate suboptimal adherence [5, 6]. Despite established efficacious treatments, 50% of diagnosed individuals discontinue clinical engagement within the first year. Among those retaining medical supervision, merely half achieve *≥*80% adherence to prescribed pharmacotherapy [7, 9]. Critically, inadequate adherence is the foremost determinant of uncontrolled HTN [8].

Given the profound economic burden imposed by poorly managed HTN—impacting healthcare systems, employers, and societal productivity—enhancing therapeutic adherence represents a pivotal opportunity for improving both clinical outcomes and resource allocation across the Americas [12]. This review examines the multifactorial determinants of non-adherence and proposes evidence-based interventions to mitigate this public health crisis.

## 2 Methods

### Study Design and Setting

A descriptive, observational, cross-sectional, and prospective study was conducted to evaluate antihypertensive therapeutic adherence among adults diagnosed with arterial hypertension. Participants were recruited from outpatient services at the General Hospital of Zone and Family Medicine Clinic No. 21 (HGZMF No. 21) of the Mexican Social Security Institute (IMSS). The investigation employed a concurrent mixed-methods approach, integrating quantitative and qualitative methodologies to comprehensively characterize adherence patterns. Study procedures adhered to STROBE guidelines for observational research.

### Participants and Sampling

The target population comprised adults (*≥*18 years) with clinically confirmed hypertension under pharmacological treatment at HGZMF No. 21. A non-probabilistic sampling strategy was implemented, consecutively recruiting eligible patients during routine consultations until reaching the predetermined sample size. Inclusion criteria required: 1) verified hypertension diagnosis, 2) active antihypertensive prescription, 3) institutional beneficiary status, and 4) written informed consent. Exclusion criteria encompassed: 1) cognitive impairment (clinically assessed), 2) language barriers, or 3) comorbidities precluding reliable participation.

Sample size was calculated using the finite population formula:

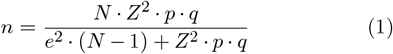

where:

- *N* = Estimated hypertensive population (1,200 based on institutional records)
- *Z* = 1.96 (95% confidence level)
- *p* = Expected adherence prevalence (55% = 0.55)
- *q* = 1 − *p* = 0.45
- *e* = Margin of error (0.085)

### Calculation

*Numerator* :

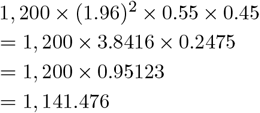

*Denominator* :

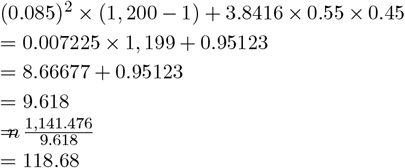

### Attrition Adjustment (10%)

Considering a 10% potential non-response rate:

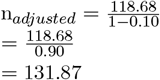

*This, the final adjusted sample size was rounded up to 132 participants*.

This yielded an initial target of 131 participants. After accounting for two unsuccessful questionnaire completions (one due to acute clinical deterioration and one withdrawal during interview), the final analyzed sample comprised 129 participants. Stratification was performed across six age cohorts (18-29, 30-39, 40-49, 50-59, 60-69, *≥*70 years) using proportional allocation based on institutional demographic data.

Data were gathered through structured face-to-face interviews using a validated questionnaire assessing sociodemographic data, clinical parameters, and therapeutic adherence (total, partial, or non-adherence). Trained personnel conducted interviews in private clinical settings from October 2024 to March 2025, averaging 22 minutes each. Data were entered and managed in Excel, analyzed, and visualized with Jamovi. A double-entry system ensured data accuracy. Results are presented descriptively using graphs, without inferential statistics. Manuscript preparation was done using LaTeX Overleaf.

Manuscript preparation, including formatting and figure integration, was facilitated by LaTeX Overleaf.

### Ethical Considerations

The study protocol received approval from the IMSS Research Ethics Committee (Approval No. R-2024-10-015) on 15 October 2024. The investigation complied rigorously with Declaration of Helsinki principles and Mexican regulatory standards (NOM-012-SSA3-2012). Written informed consent was obtained from all participants after comprehensive explanation of study objectives. Anonymization protocols included replacement of identifiers with unique codes, secure data encryption, and restricted database access. Participants retained unconditional withdrawal rights without care compromise. Original research databases remain archived under institutional custody with regulated access provisions.

## 3 Results

The study population comprised a total of 129 participants. With respect to sex distribution, 69 participants (53.5%) were female and 60 (46.5%) were male (Figures 2 and 3).

**Figure 1.**
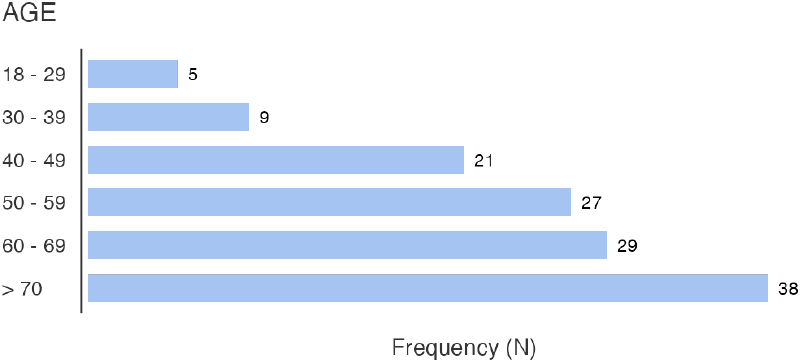
Age group distribution of study participants

**Figure 2.**
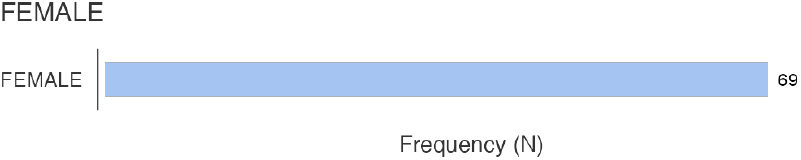
Distribution of female participants by age group

**Figure 3.**
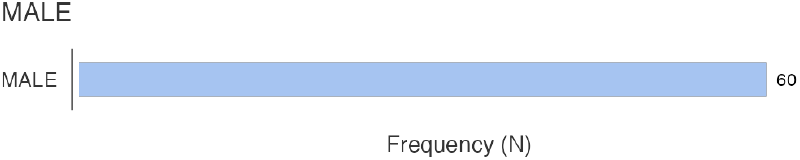
Distribution of male participants by age group

Age distribution analysis (Figure 1) demonstrated a progressive increase in participant frequency across ascending age categories. The most represented age group was 70 years (n = 38), followed by participants aged 60–69 years (n = 29) and 50–59 years (n = 27). Intermediate frequencies were observed in the 40–49 (n = 21) and 30–39 (n = 9) age groups. The lowest representation corresponded to the 18–29-year category, which included only five participants.

### Clinical Characteristics of the Study Population

Analysis of comorbid conditions within the study population (Figure 4) revealed that diabetes was the most prevalent, affecting 45 participants (34.9%). Cardiac diseases were reported with equal frequency (n = 45, 34.9%), followed by renal disease in 12 individuals (9.3%). Other reported conditions included overweight and obesity (n = 9, 7.0%), respiratory diseases (n = 6, 4.7%), and a variety of other disorders grouped under the category “Other diseases” (n = 11, 8.5%). Notably, 34 participants (26.4%) reported no chronic conditions at the time of assessment.

**Figure 4.**
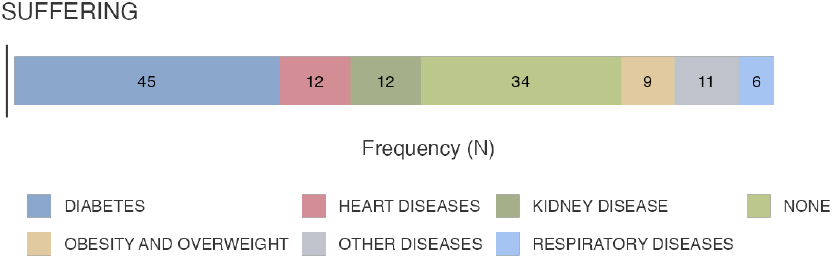
Comorbidities present in the study population

These comorbidities represent critical determinants of clinical complexity and therapeutic adherence. The elevated prevalence of diabetes and cardiovascular disease highlights the necessity for multidisciplinary, integrated care models capable of addressing coexisting chronic conditions. Although less frequent, renal disease, obesity, and respiratory illnesses contribute substantially to the overall burden of morbidity, reinforcing the need for comprehensive patient management strategies. A clear understanding of the distribution and clinical implications of these comorbidities is essential to guide tailored interventions aimed at improving both adherence and long-term health outcomes.

Regarding time since diagnosis (Figure 5), the majority of participants (n = 56) had been living with their condition(s) for more than 10 years. This was followed by 36 participants diagnosed 5–10 years prior, and 26 participants with a diagnosis between 1 and 4 years. The least represented group consisted of individuals diagnosed within the past year (n = 11).

**Figure 5.**
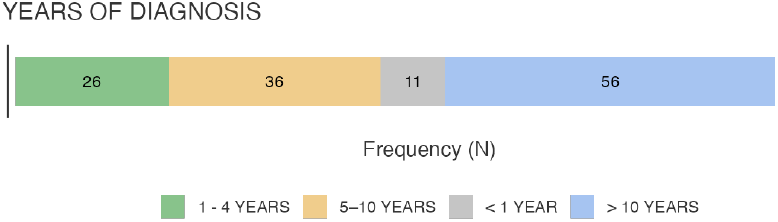
Time since initial hypertension diagnosis

### Correlation Analysis Between Age and Time Since Diagnosis

A stratified analysis of age groups in relation to the duration since diagnosis (Figure 6) revealed distinct distribution patterns suggestive of an age-related trend in chronic disease progression.

**Figure 6.**
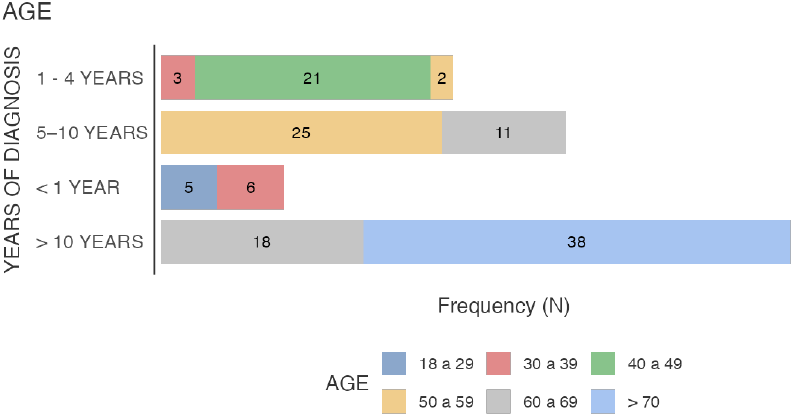
Correlation between age groups and time since diagnosis

Among participants diagnosed within the past year (n = 11), all cases were concentrated in the younger age brackets: 18–29 years (n = 5) and 30–39 years (n = 6). In the group diagnosed 1–4 years prior (n = 26), the highest frequency corresponded to the 40–49-year cohort (n = 21), followed by a smaller representation in the 30–39 (n = 3) and 50–59 (n = 2) age groups.

For participants with a diagnosis dating back 5–10 years (n = 36), most were aged 50–59 years (n = 25), with the remainder falling within the 60–69-year group (n = 11). Finally, among those with a disease duration exceeding 10 years (n = 56), all individuals were from older age categories: 18 were aged 60–69 years, and the majority (n = 38) were over 70 years of age.

These findings suggest a clear positive correlation between increasing age and the chronicity of diagnosed conditions. The progressive shift of longer disease duration toward older cohorts underscores the cumulative burden of chronic illness over time and highlights the importance of age-specific care strategies to address the evolving needs of aging populations.

### Pharmacotherapy and Therapeutic Adherence Analysis

The analysis of antihypertensive pharmacotherapy among participants (Figure 7) identified Losartan as the most frequently prescribed agent (n = 33), with its use spanning a broad age range, from 18 to 69 years. A notable concentration of Hydrochlorothiazide users (n = 18) was observed within the 50–69-year age groups, while Metoprolol (n = 12) was predominantly used among individuals aged 60–69 years. Amlodipine (n = 11) was primarily prescribed to participants aged 60 years and older, including 7 individuals aged 60–69 and 4 over 70 years.

**Figure 7.**
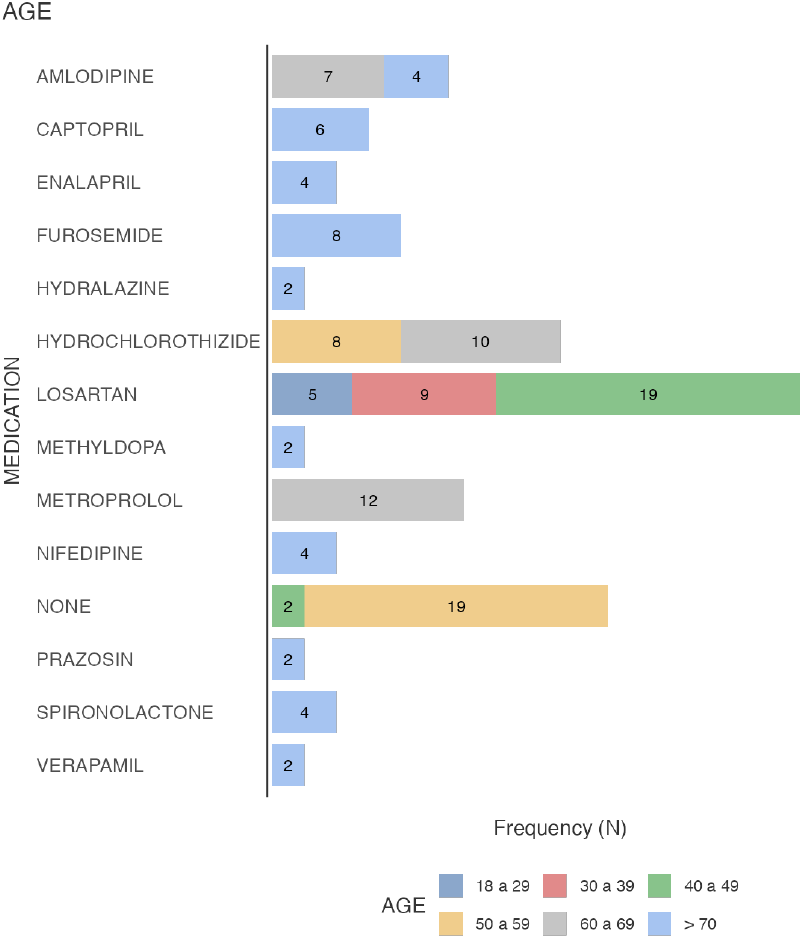
Antihypertensive pharmacotherapy distribution by age group

Medications such as Captopril (n = 6), Enalapril (n = 4), Furosemide (n = 8), Hydralazine (n = 2), Methyldopa (n = 2), Nifedipine (n = 4), Prazosin (n = 2), Spironolactone (n = 4), and Verapamil (n = 2) were reported almost exclusively among participants over 70 years of age, suggesting a trend toward polypharmacy and more complex antihypertensive regimens in older adults.

Interestingly, a subgroup of 21 participants reported no current use of antihypertensive medication, most of whom were in the 50–59-year age range (n = 19), with a smaller subset in the 40–49 group (n = 2). This finding may reflect early-stage disease, treatment discontinuation, or barriers to access or adherence.

With respect to therapeutic adherence (Figures 8 and 9), the total sample (N = 129) was categorized into three adherence levels: partial, non-adherent, and fully adherent. Partial adherence represented the majority (n = 105), with the most commonly involved medications being Verapamil (n = 21), Hydrochlorothiazide (n = 18), and Losartan (n = 18).

**Figure 8.**
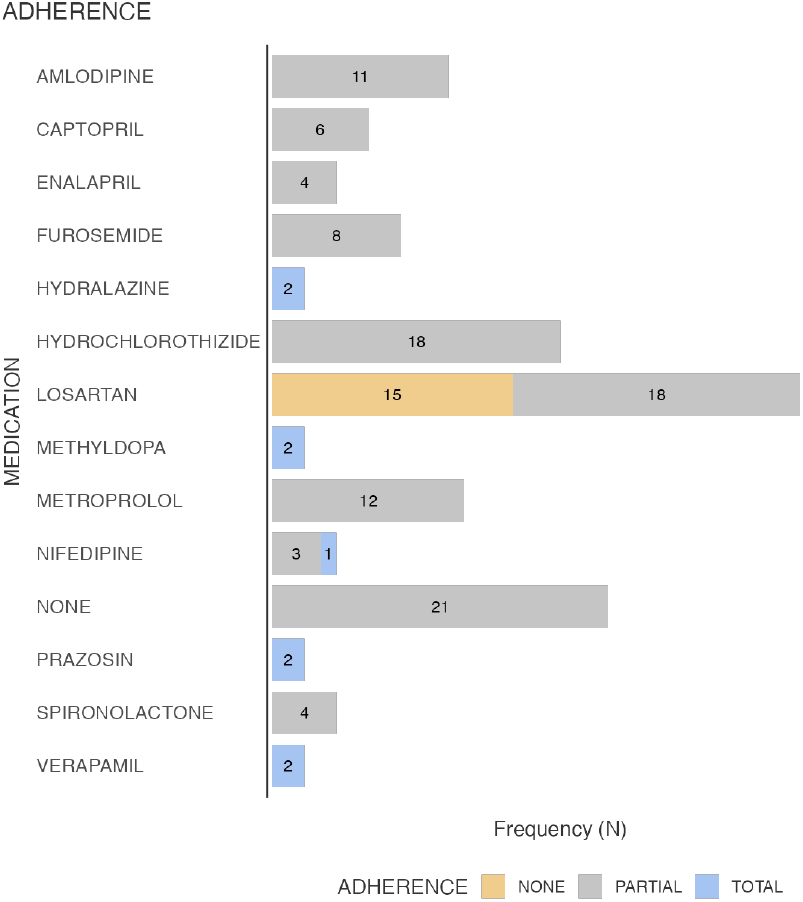
Antihypertensive pharmacotherapy distribution by adherence distribution

**Figure 9.**
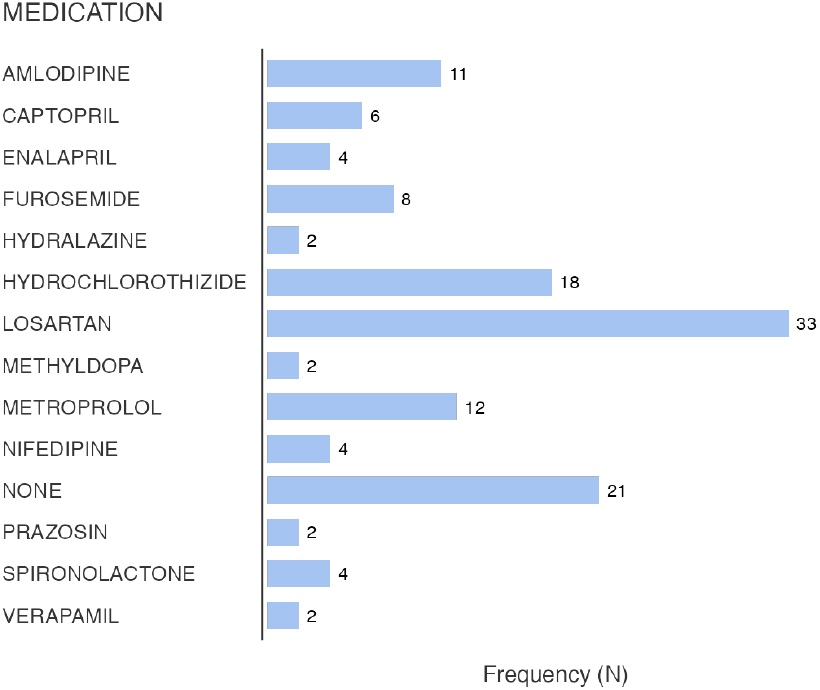
Prescription frequency of antihypertensive medications

A total of 15 participants were classified as non-adherent, all of whom were prescribed Losartan, suggesting possible challenges specific to this pharmacological regimen, such as perception of side effects, complexity of dosage, or lack of symptom control.

Complete adherence was observed in only 9 participants, distributed across the use of Amlodipine (n = 2), Captopril (n = 2), Enalapril (n = 1), Hydrochlorothiazide (n = 2), and Losartan (n = 2). This limited representation underscores the need for improved strategies to support medication adherence across all age groups and treatment regimens.

A more granular analysis of therapeutic adherence by specific antihypertensive agents (Figures 8 and 9) provided further insights into medication-specific patterns of compliance.

Partial adherence was the most frequently observed pattern across nearly all pharmacological classes. Notably, amlodipine was associated with partial adherence in 11 participants, followed by captopril (n = 6), enalapril (n = 4), furosemide (n = 8), hydrochlorothiazide (n = 18), metoprolol (n = 12), spironolactone (n = 4), nifedipine (n = 3), and losartan (n = 18). Additionally, 21 participants with no pharmacological therapy reported partial adherence to nonpharmacological recommendations (e.g., lifestyle changes), highlighting a possible divergence between clinical prescription and behavioral adherence.

In contrast, non-adherence was observed exclusively among losartan users (n = 15), suggesting this agent may be particularly susceptible to poor compliance, possibly due to side effects, cost, or perceived lack of efficacy. This highlights the need for targeted interventions to address barriers specific to this medication.

Complete adherence was comparatively rare and restricted to a few agents. It was observed in two participants each using hydralazine, methyldopa, and prazosin; one participant using nifedipine; and two using losartan, as well as two individuals with hydrochlorothiazide.

These findings underscore significant variability in adherence profiles across antihypertensive medications. They emphasize the importance of personalized adherence support strategies, particularly for agents with lower adherence rates, such as losartan. Furthermore, the low overall rate of full adherence within the sample indicates a critical gap in long-term treatment effectiveness, which may contribute to suboptimal blood pressure control and increased cardiovascular risk.

### Distribution of Medication Prescription Frequency

Figure 10 illustrates the distribution of prescription frequencies for antihypertensive medications among the study participants. Losartan was the most frequently prescribed agent (n = 33), followed by a substantial number of individuals who were not receiving any listed antihypertensive medications at the time of the study (n = 21). Hydrochlorothiazide also demonstrated high utilization (n = 18), while amlodipine (n = 11), metoprolol (n = 12), furosemide (n = 8), nifedipine (n = 4), and captopril (n = 6) exhibited moderate prescription frequencies.

**Figure 10.**
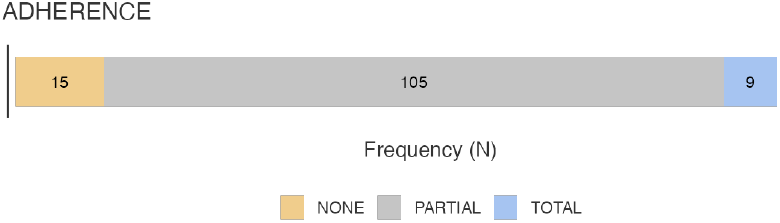
Overall therapeutic adherence distribution

Medications with the lowest prescription frequencies included hydralazine (n = 2), methyldopa (n = 2), spironolactone (n = 4), prazosin (n = 2), and verapamil (n = 2). The particularly low usage of hydralazine, methyldopa, prazosin, and verapamil suggests these agents were reserved for specific clinical indications or used in more complex therapeutic regimens, possibly within the oldest age group or in cases of treatment resistance.

### Overall Adherence Frequency

Figure 10 presents the distribution of therapeutic adherence levels within the study cohort. Partial adherence emerged as the predominant category, encompassing 105 individuals and representing the largest proportion of the sample. In contrast, complete adherence was observed in only 9 participants, indicating a relatively limited occurrence of full compliance with prescribed treatment regimens.

A total of 15 participants were classified as non-adherent, reflecting a significant subgroup for whom therapeutic guidelines were not followed consistently or at all. These findings underscore a concerning trend: while partial adherence ap-pears to be the norm, complete adherence remains uncommon, and a notable proportion of individuals are entirely disengaged from prescribed treatment. This highlights the persistent challenges in promoting optimal adherence among patients with chronic conditions and suggests the need for targeted interventions to improve compliance and health outcomes.

### Adherence Stratified by Years Since Diagnosis

Figure 11 illustrates therapeutic adherence stratified by duration since diagnosis, categorized under *Years Since Diagnosis*. A clear trend was observed wherein partial adherence increased progressively with longer disease duration. The highest frequency of partial adherence was found among participants diagnosed more than 10 years ago (n = 47), followed by those diagnosed 5–10 years prior (n = 36) and 1–4 years prior (n = 22). Notably, no cases of partial adherence were reported in the group diagnosed less than one year ago, where all 11 participants were classified as non-adherent.

**Figure 11.**
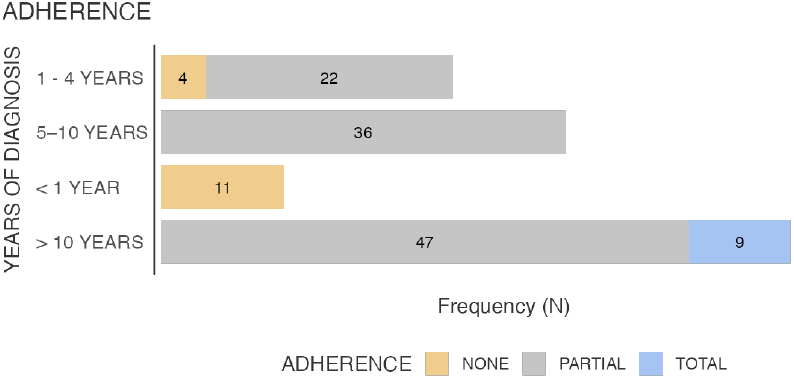
Therapeutic adherence by time since diagnosis

Complete adherence was observed exclusively among individuals with over 10 years since diagnosis (n = 9), suggesting that extended exposure to disease management may promote better adherence behaviors over time. In contrast, non-adherence was most prevalent among those diagnosed 5–10 years ago (n = 11), followed by a smaller number in the 1–4 years group (n = 4). No cases of non-adherence were identified in the 1 year or 10 years groups.

These patterns indicate a possible U-shaped relationship between time since diagnosis and adherence: non-adherence appears to peak during intermediate stages, while both recent and long-standing diagnoses show higher rates of either non- or full adherence, respectively. These findings emphasize the need for continuous adherence support, particularly during the mid-term phase of disease progression, where risk of disengagement may be elevated.

Taken together, these trends highlight the importance of longitudinal adherence support, particularly targeting the mid-term phase of disease, when patients may become complacent or disillusioned. Future interventions should aim to reinforce motivation, improve therapeutic literacy, and offer personalized adherence counseling throughout the disease continuum.

### Adherence Stratified by Age Groups

Figure 12 displays the distribution of therapeutic adherence across different age groups, revealing age-related patterns in treatment behavior.

**Figure 12.**
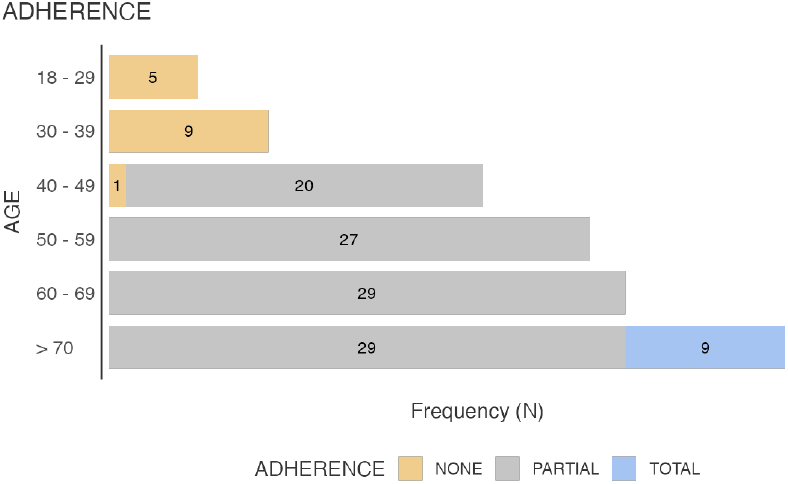
Therapeutic adherence by age groups

Complete adherence was observed exclusively among participants aged over 70 years. All nine individuals in this group demonstrated full adherence to their prescribed treatment regimens. This finding suggests that older adults may exhibit stronger treatment commitment, possibly due to increased health awareness, more frequent healthcare engagement, or longer exposure to chronic disease management strategies. No cases of complete adherence were reported in any younger age category.

Partial adherence was predominantly found among middleaged and older adults. The 60–69-year group accounted for 29 cases, followed by 27 in the 50–59 group, and 20 in the 40–49 group. These results suggest that although complete adherence may be less common in these age ranges, a substantial proportion of individuals maintain some level of treatment consistency. This pattern may reflect an intermediate stage of behavioral adaptation to chronic illness and its therapeutic demands.

Non-adherence was confined entirely to younger age groups. All participants aged 18–29 years (n = 5) and 30–39 years (n = 9) reported complete non-adherence, as did one individual in the 40–49 age group. Notably, no cases of non-adherence were observed among participants aged 50 years or older. This suggests a pronounced inverse relationship between age and treatment non-adherence, with younger cohorts being more likely to disengage from prescribed care. The findings imply that adherence may improve over time as individuals mature and accumulate experience in managing their condition.

## 4 Discussion

This cross-sectional analysis of therapeutic adherence among hypertensive patients treated within Mexico’s public healthcare system revealed a concerning trend: 62% of participants demonstrated suboptimal adherence, with partial adherence as the predominant category. Stratification by age uncovered a marked inverse adherence gradient, wherein younger adults (18–29 years) exhibited the highest rates of non-adherence, while older populations, particularly those aged 70 years and above, demonstrated significantly better adherence outcomes, including complete adherence in all individuals within this group. Notably, no instances of complete adherence were recorded among any of the younger age cohorts.

This pattern, which contrasts with traditional adherence models in high-income countries—where older adults are often presumed to face greater adherence challenges due to polypharmacy or cognitive decline—may be partially explained by Mexico’s cultural and healthcare context. Specifically, collectivist family structures, in which elderly individuals often reside with relatives, may facilitate medication oversight, routine reinforcement, and emotional support. In contrast, younger individuals, especially those entering the workforce or higher education, may face greater autonomy, lifestyle disruptions, and weaker perceptions of vulnerability, contributing to poorer adherence behavior.

Interestingly, adherence also appeared to improve with increased duration since diagnosis. Among participants with over 10 years of diagnosed hypertension, both partial (n = 47) and complete adherence (n = 9) were most prevalent, while non-adherence was entirely absent in this group. Conversely, individuals diagnosed less than one year ago were exclusively classified as non-adherent (n = 11). This U-shaped association suggests that therapeutic adherence may initially be hindered by denial, lack of routine, or insufficient patient education, but that, over time, disease acceptance, treatment habituation, and self-management skills tend to improve.

Importantly, the highest rate of non-adherence was observed in patients 5 to 10 years post-diagnosis, a stage that may correspond to therapeutic fatigue, where patients feel asymptomatic, overburdened by chronic care demands, or disillusioned with treatment effectiveness. This finding underscores the need for ongoing support during the mid-stage of chronic disease management, rather than relying solely on early education or late-stage care.

From a structural perspective, institutional barriers were also implicated in non-adherence. Specifically, 28% of non-adherent participants cited medication unavailability at IMSS pharmacies, pointing to supply chain failures that undermine individual efforts and physician prescriptions alike. Addressing these logistical gaps is critical to realizing the full benefit of pharmacological strategies at the population level.

These findings carry several important implications for public health policy and intervention design in Mexico and comparable settings.

First, age-specific adherence interventions are urgently needed. For younger adults, strategies should focus on mobile health tools, gamification of adherence, workplace-based education, and culturally tailored campaigns that address risk perception and lifestyle compatibility. Conversely, older adults may benefit more from caregiver training, simplified regimens, and polypharmacy reviews.

Second, mid-diagnosis-stage patients (5 to 10 years) represent a high-risk group for disengagement. Adherence support programs should integrate motivational interviewing, peer support groups, and digital monitoring tools targeting this specific window of vulnerability.

Third, at the health system level, reforms should prioritize stable medication supply chains, especially within IMSS and ISSSTE systems, ensuring that medication unavailability does not become a structural barrier to adherence. Strengthening pharmacy logistics, real-time inventory systems, and inter-institutional procurement may mitigate these lapses.

Methodologically, this study is strengthened by its use of validated adherence measures, age-stratified and time-since-diagnosis analysis, and inclusion of patients from real-world public healthcare settings. However, its cross-sectional nature limits causal inference, and the absence of biochemi-cal adherence verification (e.g., pill counts or blood pressure control metrics) may underestimate or misclassify adherence status. Additionally, factors such as health literacy, comorbidities, depression, and health system navigation skills were not explored but may significantly influence adherence behaviors.

## 5 Conclusion

This study revealed substantial therapeutic adherence deficiencies among hypertensive patients managed within Mexican Institute Social Sucurity, with 62% of the population demonstrating suboptimal adherence. The findings highlight a pronounced age gradient, wherein complete adherence was exclusively observed in older adults (70 years), while younger cohorts (18–39 years) exhibited disproportionately high levels of non-adherence. Additionally, longer disease duration (10 years since diagnosis) was associated with improved adherence, suggesting that sustained exposure to treatment may facilitate behavioral adaptation and self-management skills.

These patterns underscore the need for age-sensitive and temporally calibrated interventions that not only address individual clinical variables—such as regimen complexity and adverse effects—but also incorporate sociocultural dimensions, including perceived vulnerability, familial dynamics, and health literacy. Moreover, systemic barriers such as medication shortages, fragmented care, and prolonged wait times remain critical obstacles to sustained adherence and must be addressed through institutional reforms.

To effectively reduce the burden of uncontrolled hypertension and its cardiovascular sequelae, multifaceted strategies are required. These should include the implementation of structured patient education programs, digital adherence monitoring tools, targeted outreach for younger adults, and expanded access to essential medications through pharmacy system optimization. The integration of these elements into Mexico’s national health policies is essential for improving adherence outcomes and ensuring the long-term sustainability of hypertension control efforts within the IMSS-insured population.

## Acknowledgements

The authors express profound gratitude to the nursing and administrative staff of HGZMF No. 21 for their invaluable support in participant recruitment and data collection. We acknowledge the statistical guidance provided by the Department of Medical Education and Research. Most im-portantly, we thank the study participants for generously sharing their time and experiences.

## Funding

This research did not receive any specific grant from funding agencies in the public, commercial, or not-for-profit sectors. The study was self-funded by the authors. Infrastructure support was provided by the General Hospital of Zone and Family Medicine Clinic No. 21, IMSS Léon, Guanajuato, Mexico

## Authors’ Contributions

All authors contributed to the development of this study, including its conceptualization, methodology, investigation, and writing. All authors reviewed and approved the final version of the manuscript and agree to be accountable for all aspects of the work.

## Conflict of Interest

The authors declare no conflicts of interest. No financial or personal relationships influenced this work.

## Data Availability

The datasets generated and analyzed during this study are not publicly available due to institutional privacy policies. However, they may be made available from the corresponding author upon reasonable request, subject to approval by the Instituto Mexicano del Seguro Social (IMSS) data sharing agreements. Interested parties should contact the corresponding author for further information.

## Ethical Statement

The study protocol was approved by the IMSS Research Ethics Committee. All procedures followed the ethical standards of the Declaration of Helsinki and Mexican regulatory standards (NOM-012-SSA3-2012). Written informed consent was obtained from all participants.

## Declaration of AI Usage

During the preparation of this manuscript, no artificial intelligence tools, were used at any stage of its development. All scientific content, data interpretation, language refinement, and conclusions are entirely the result of human intellectual work. The manuscript was prepared and typeset using LaTeX Overleaf.

## Notes

### Competing Interest Statement

The authors have declared no competing interest.

### Author Declarations

Ethical approval for this study was granted by the Comite Local de Etica en Investigacion en Salud del Hospital General de Zona con Medicina Familiar No. 21, Instituto Mexicano del Seguro Social (IMSS), Leon, Guanajuato, Mexico. The committee reviewed and approved the research protocol under institutional guidelines for studies involving human subjects.

